# Radial and Axial Fractional Shortening for Rapid Estimation of Left Ventricular Ejection Fraction: A Computational Analysis

**DOI:** 10.1101/2023.06.22.23291770

**Authors:** Robert D. Rifkin, Michael W. Rich

**Affiliations:** Washington University School of Medicine St. Louis, MO; From the Division of Cardiology, Department of Medicine Washington University School of Medicine, St. Louis, Missouri

**Author notes:** **Conflicts of Interest: None**. Address for correspondence: Robert D. Rifkin, M.D.

## Abstract

**BACKGROUND AND PURPOSE:** Simpson Rule based planimetry remains the gold standard for left ventricular (LV) ejection fraction (EF) but due to sub-optimal endocardial delineation, planimetry is not feasible in many cases. The purpose of this study was to derive and to analytically evaluate the theoretical accuracy of several simple novel formulas for estimating EF in ventricles with uniform wall motion using only the radial diameter and axial length LV fractional shortening (F_D_ and F_L_ respectively), which are less subject to image quality limitations than planimetry.

**METHODS:** A truncated ellipsoidal model of the LV at end-diastole and end-systole was assumed leading to a novel compact formula for directly calculating an exact EF that is identical to the Simpson Rule EF when the ventricle has ellipsoidal end-diastolic (ED) and end-systolic (ES) shapes.

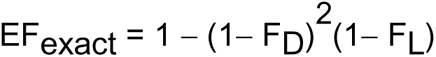

Three linear formulas were then developed to directly calculate an approximation to the exact EF without intermediate calculation of volumes. To avoid population selection bias in this initial investigation the linear coefficients were determined by an analytic graphical optimization procedure that minimized the EF errors compared to the exact EF over the full range of exact EFs from 0 to 80% (F_D_ and F_L_ in percent).

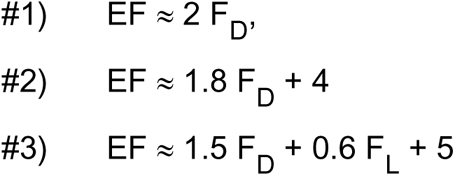

**RESULTS:** In single factor linear approximations #1 and #2, the EF difference from the exact EF was dependent on the ratio F_L_/ F_D_, which is normally between 40 and 50%. Subject to the assumption of ellipsoidal LV shape at ED and ES, for ratios of F_L_/ F_D_ between 20% and 80%, and over EFs ranging from 0 to 55%, the calculated differences from the exact EF were less than 6 EF points. At extremes of the ratio, less than 20% or over 80%, or for EFs over 55%, the difference could reach or exceed 10 points. Formula #2 produced slightly smaller differences than formula #1. Formula #3 reduced the difference error substantially to less than 4 EF percentage points regardless of the F_L_/ F_D_ ratio except at EFs over 75%.

**CONCLUSIONS:** In uniformly contracting ventricles, simple linear formulas could provide rapid estimation of EF and may be helpful when image quality degrades planimetric EF accuracy, as well as in point-of-care echocardiography where planimetry is not feasible. Validation of these formulas through empirical testing is warranted.

## INTRODUCTION

Simpson Rule based planimetry remains the gold standard for optimal echocardiographic measurement of left ventricular (LV) ejection fraction (EF) because it can be applied in ventricles of any shape at end-diastole (ED) and end-systole (ES) [1]. However, due to sub-optimal image quality, planimetry is often not feasible or is of reduced accuracy. Even when wall motion is visualized adequately in moving image loops, when frozen at ED and especially ES, it is often difficult to confidently identify endocardium for planimetry. This has led to readers utilizing overall “eye ball” visual estimates of LV EF from the video loops without any quantitative measurement [2]. The use of sonographic contrast has mitigated the problem substantially, but its use is limited primarily to inpatient laboratories and it is rarely used in the outpatient setting. Accordingly, a variety of alternative methods for assessing LV function have evolved. Visual based segmental scoring schemes have been used as a semi-quantitative surrogate for EF when planimetry is not feasible and segmental wall motion abnormalities (SWMA) are present [3,4]. However, these alternatives are difficult to apply in cases of uniform LV dysfunction in the absence of a normally contracting segment to use as a visual benchmark. Uniformly or nearly uniformly contracting ventricles represent a large group of clinical cases comprising most forms of non-ischemic cardiomyopathy including hypertensive heart disease, toxic myopathy and myocarditis. A variety of other indices of LV performance have been suggested for use in technically difficult cases [5], but none of these indices is easy to use and none report an EF directly.

The potential for accurate measurement of EF in cases of uniform LV contraction using the radial diameter (F_D_) and axial length (F_L_) fractional shortening of the LV has not been systematically analyzed. Radial fractional shortening was originally used alone to estimate EF by the Cube and Teicholz methods [6,7], but these were tedious to use, involving intermediate calculation of ED and ES volumes, and required a computer.

The purpose of this investigation was: (1) to develop simple formulas that do not require a computer system and can be applied in cases without significant SWMA to obtain the approximate EF without intermediate calculation of LV volumes; and (2) to examine their accuracy in a geometric numerical simulation of LV contraction.

These formulas could be useful to clinicians for estimating EF both in the echocardiography laboratory, especially in the community where contrast is rarely employed, as well as in point of care (POC) echocardiography, the use of which is increasing.

## METHODS

### Exact EF in Ellipsoidally Shaped LV

A novel compact formula for converting radial and axial LV fractional shortening, F_D_ and F_L_ respectively, defined as (ED dimension – ES dimension)/(ED dimension), directly into LV EF was derived from basic geometric considerations that provides an exact EF in chambers that have ellipsoidal ED and ES shape where F_D_ and F_L_ are expressed as fractions (see appendix 1):

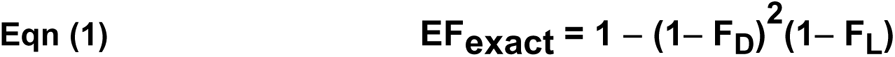

### Relation between F_D_ and F_L_

Normal mean values for radial and axial fraction shortening vary in the literature but typical values are F_D_ between 30% and 35% and F_L_ between 15% and 20%, respectively [8,9]. Thus, in ventricles contracting normally, axial shortening fraction is approximately 40% - 60% of the radial fractional shortening. When LV function deviates from normal this ratio may change but except for rare circumstances, axial shortening typically remains less than radial shortening fraction. Expressing the formula for exact EF in terms of this ratio provides simplification and geometric insight in the analysis. We defined this ratio as:

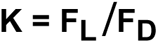

Substituting F_L_ = KF_D_ into Eqn 1 leads to an alternative form of Equation 1 for the exact EF in an elliptically shaped model:

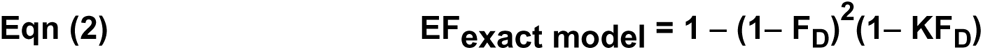

As a first step to obtaining linear formulas, we expanded the squared term, (1− F_D_)^2^ to obtain this algebraically equivalent form of the exact equation (appendix 3):

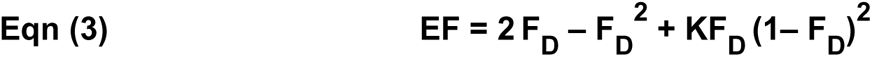

Note that the 2^nd^ and 3^rd^ terms are squares of fractions and are therefore both smaller than the 1^st^ term, 2F_D_. Importantly, since they are also of opposite sign, they tend to cancel each other and can be ignored leaving only the 1^st^ term,_2 F_D_ as an approximation to the EF, or EF ≈ 2 F_D_

As an illustration, typical normal LV radial fractional shortening is 33% or F_D_ = 0.33 and axial fractional shortening is 20% or F_L_ = 0.2. The second term of the expansion is **−**0.109, the third term is +0.089. Their sum is **−**0.019, which is much smaller than the main term 2 F_D_= 0.66. Thus, the approximate EF, 2 F_D_, is 2 x .33 or 0.66 and is very close to the EF = 0.64 given by the complete Exact formula including all terms. We designate this approximation as Model 1:

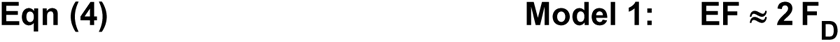

In Model 1, the LV EF is estimated by simply doubling the LV radial fractional shortening, F_D_.

In the following analysis, the accuracy of this approximation and other linear formulas that include both the axial and radial fractional shortening will be examined over the entire spectrum of physiologic EFs and fractional shortening values by comparison to the exact formula, Eqns 1 and 2.

### General Linear Models

We generalized Model 1 to include the axial LV fractional shortening as follows:

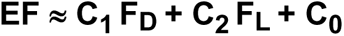

In Model 1, C_1_ = 2 and C_2_ and C_0_.are both zero. The exact formula (equations 1 and 2) were used in a geometric numerical simulation of LV contraction. The linear formula coefficients were adjusted or “optimized” to minimize the difference between the linear formula EF and the gold standard EF provided by the exact formula for clinically plausible values of F_D_ and F_L_.

### Model 2

To better reflect variations in the 2^nd^ and 3^rd^ terms of equation 3 and maintain model accuracy over a wider range of fractional shortening values, two terms are included in this one-factor model: EF ≈ C_1_F_D_ + C_0_. The value of C_1_ was chosen so that, if C_0_ = 0, the model is a straight line passing through an EF of 0% when F_D_ = 0 and a normal EF of 64% when F_D_ = 35% (i.e. Estimated EF/Normal EF = Measured F_D_/Normal F_D_). Thus, if C_0_ = 0, EF = (F_D_/35) 64 = 1.83 F_D_ so the EF is estimated by a line of slope 1.83 passing through EF =0 and EF = 64%. To simplify, C_1_ is approximated as 1.8. Thus Model 2 is:

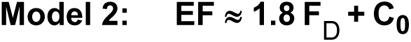

where the value of C_0_ will be determined to achieve the closest agreement with the exact EF, equation 1, over the full range of clinical EFs by an optimization procedure described below.

### Model 3

To more accurately accommodate disparities between the magnitude of radial and axial fractional shortening, a two-factor model including both shortening components was investigated:

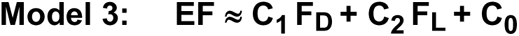

As in models 1 and 2, the value of the constants will be determined to achieve the closest agreement with the exact EF, equation 1, by an optimization procedure as described below.

### Computational optimization of linear formula coefficients by numerical simulation of contraction (figure 1)

The exact formula provides the “gold standard” EF based upon a geometric model of the LV shaped as a prolate ellipsoid at end diastole and end systole. To derive a specific set of coefficients for each of our three linear formulas, we calculated the percentage point difference between the linear model’s EFs and the exact EF given by equation 2 for EFs from 0 to 75%. These differences were graphed on the Y-axis as “EF ERROR” (or ΔEF) as a function of the exact EF on the X-axis for EFs from 0% to 75% for a particular value of K (figure 1). To take into consideration independent variations in F_D_ and F_L_, a family of seven curves was created for each model for seven values of K: 0%, 20%, 40%, 60%, 80%, 100% or 120%. Wherever an error curve crosses the X-axis the EF error of the model is zero. The distance from each curve for a particular value of K to the X-axis at a given EF on the X-axis gives the formula’s error ΔEF that is shown on the Y-axis. Thus, for each value of K and for exact EFs from 0% to 75%, we calculated:

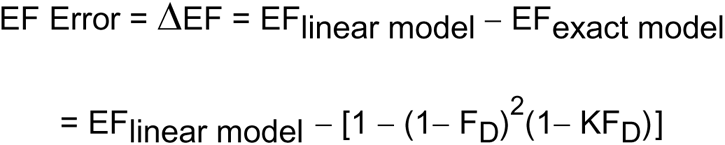

**Figure 1:**
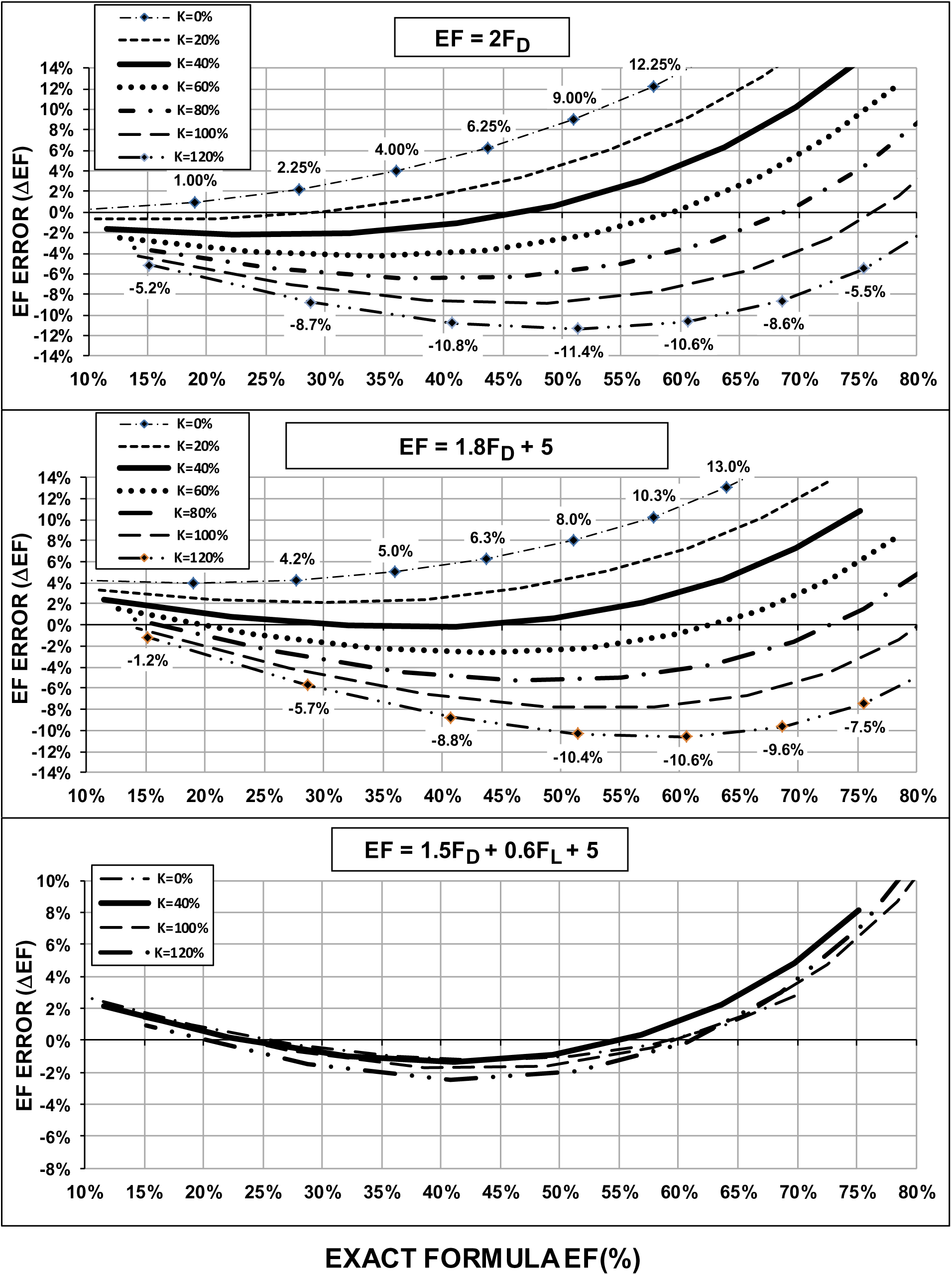
Theoretical curves of EF error in percentage points for three geometrically optimized linear approximations to the exact EF given by the structural model: EF_exact_ = 1 − (1− F_D_)^2^(1− F_L_). Separate curves are shown for 7 values of the ratio of axial to radial shortening, K = F_L /_ F_D_. The coefficients in models 2 and 3 (middle and bottom panels) have been adjusted to minimize the EF error for the normal values of K between 40 and 50% over the clinically most important range of EF values from 0 to 65%, as reflected by aligning the heavy black line as close to the X-axis as possible over this range.

The procedure for construction of each curve in Figure 1 is illustrated in Table 1 for the case of EF ≈ 1.8 F_D_ + 4 with K = 60%. To generate the exact EF values for a particular value of K, we varied F_D_ from 5% to 50% for that value of K in exact EF equation 2. These assumed values of radial fractional shortening, F_D_, are listed in column 1 of Table 1. The corresponding axial fractional shortening value, F_L_, for each F_D_ value is found from the definition of K (assumed to be 60% in this example) by solving for F_L_ (F_L_ = KF_D_ = 0.6 F_D_). The corresponding values of F_L_ are listed in column 2 in percent. Then the exact EF, as calculated from Eqn #1 for each F_L_ and F_D_ pair listed in columns 1 and 2, is shown in column 3 (with per cents expressed as fractions for calculation). The EF by the linear formula for the given values of F_D_ and F_L_ is shown in column 4 (in this single factor formula example, F_L_ does not appear in that calculation). The difference, or EF error in the linear formula, defined as EF_linear model_ − EF_exact model_ is displayed in column 5. The curve in Figure 1 (middle graph) for this value of K and this linear formula is generated by plotting the values in column 5 on the Y axis versus the values in column 3 on the X axis. This procedure is repeated for each value of K (0%, 20%, 40%, 60%, 80%, 100% or 120%) to create the family of curves in Figure 1 for this linear formula.

**TABLE 1.**
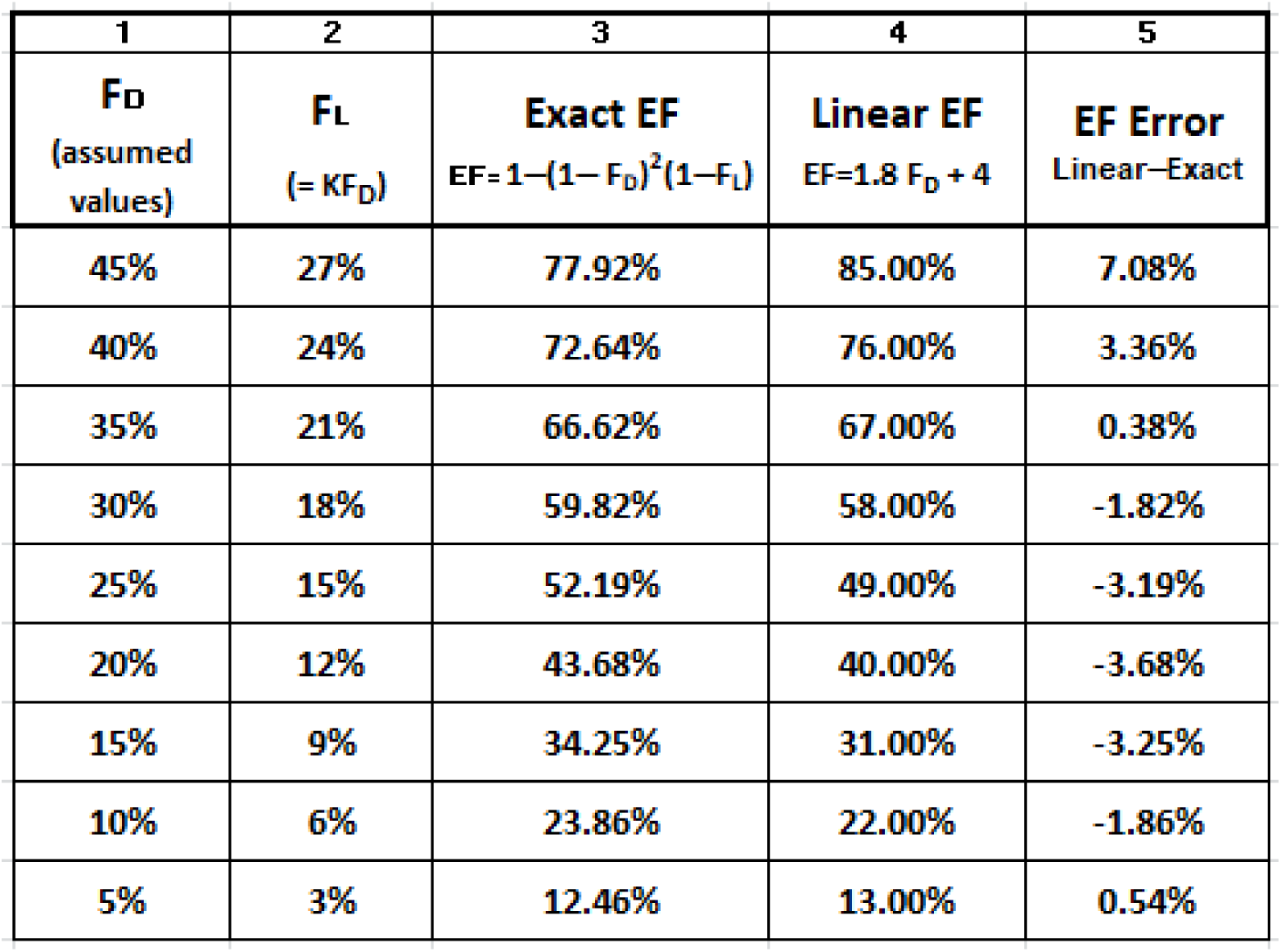
Linear EF ≈ 1.8 F_D_ + 4 with K = 60% (0.6) Values are shown in % but formulas require decimal fractions. EF Error is in percentage points of EF difference

| 1 | 2 | 3 | 4 | 5 |
| --- | --- | --- | --- | --- |
| **F<sub>D</sub>**<br>(assumed<br>values) | **F<sub>L</sub>**<br>(= K F<sub>D</sub>) | **Exact EF**<br>$EF = 1 - (1 - F_D)^2 (1 - F_L)$ | **Linear EF**<br>$EF = 1.8 F_D + 4$ | **EF Error**<br>Linear-Exact |
| 45% | 27% | 77.92% | 85.00% | 7.08% |
| 40% | 24% | 72.64% | 76.00% | 3.36% |
| 35% | 21% | 66.62% | 67.00% | 0.38% |
| 30% | 18% | 59.82% | 58.00% | -1.82% |
| 25% | 15% | 52.19% | 49.00% | -3.19% |
| 20% | 12% | 43.68% | 40.00% | -3.68% |
| 15% | 9% | 34.25% | 31.00% | -3.25% |
| 10% | 6% | 23.86% | 22.00% | -1.86% |
| 5% | 3% | 12.46% | 13.00% | 0.54% |

Using these graphical displays of EF error, we adjusted the value of the coefficients, C_0_, C_1_, and C_2_ in models 2 and 3 to visually minimize the deviation of the curve for K = 40% over the range of EFs from 0% to 65% from the X-axis, which represents zero model error,.In Model 1, C_1_ and C_0_ have already been set to values 2 and 0, respectively, thus:

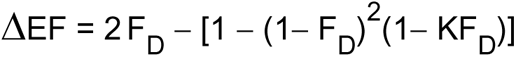

ΔEF versus the EF_exact model_ was graphed for each of the seven values of K to obtain the family of six curves (Figure 1 top panel).

For Model 2, C_1_ has been set to 1.8, thus EF = 1.8 F_D_ + C_0_. By graphing ΔEF versus EF_exact model_ and varying C_0_, we found the visually optimum value of C_0_ to be 4, resulting in the equation EF = 1.8 F_D_ + 4 (Figure 1, middle panel). Thus, graphically optimized model 2 is:

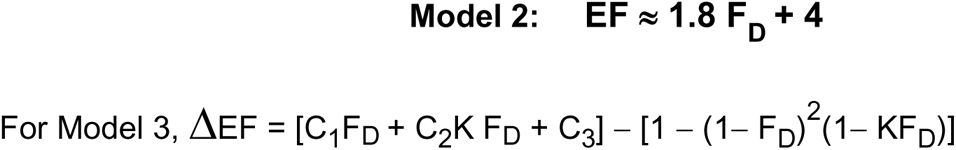

By visual inspection, we determined the optimal coefficient values for model 3 by choosing values that minimized the visual deviation of the curve for K = 40% from the zero error horizontal baseline (x-axis) in Figure 1 (bottom panel) for which the error, ΔEF = 0). These coefficients were found to be C_1_= 1.5, C_2_= 0.6, C_3_ = 5 (figure 1, bottom panel).

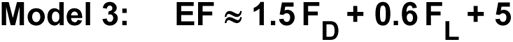

This study was approved by the Human Research Protection Office of the Institutional Review Board at Washington University in Saint Louis, Missouri.

## RESULTS

### Agreement Between Optimized Linear Models and Exact Geometric Formula EF

Figure 1 shows families of curves of the EF error expressed as the percentage point difference (on the Y-axis) from the actual exact EF (on the X-axis) for each of the three linear models:

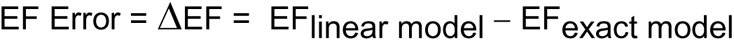

The ratio of axial to radial shortening, K = F_L_/F_D_, is a parameter in each graph ranging from 0% to 120%. As noted above, the exact model EF (eqns 1 and 2) is identical to a planimetric (“true”) EF for a chamber that maintains an ideal ellipsoidal shape at end-diastole and end-systole regardless of the proportions of axial to radial length. Graphs of the EF error (Y-axis) versus true or exact LV EF (X-axis) for constant values of K permit the relationship of the EF error to both the exact EF and to the fractional shortening ratio (K) to be easily recognized (Figure 1). In Figure 1, the curves for each value of K in each graph are, from top curve to bottom curve: K = 0%, 20%, 40%, 60%, 80%, 100%, 120%.

### Single Factor Models, #1 and #2

(top and middle graph, Figure 1): The EF error depends on the value of K as well as on the exact EF. When K remains near the normal values of 40 - 50% (heavy solid lines in graphs), error values are less than 6 percentage points over the entire range of EFs up to 65%, and the spread of error values tends to be symmetric around zero error. The EF errors increase as K deviates above or below the normal range of 40 – 50%. Thus, for K values of 20% or 100%, the error is higher, but is still less than 8 EF percentage points over the range of EFs from 0 to 55%. At more extremes of the ratio K, less than 20% or over 100%, and for EFs over 60%, error could reach or exceed 10 EF points.

It can be seen from the middle graph that errors could be reduced with the equation EF ≈ 1.8 F_D_ + 4 compared with the equation EF ≈ 2 F_D_. The spread of the curves is slightly less. The reduction in errors is evident in the values of error shown along the curves for the lowest and highest value of K for the top and middle graph.

### Two Factor Model, #3

(bottom graph, Figure 1): In model #3, which included the axial fractional shortening, error was dramatically reduced for all EFs. Only at EFs above 65% did error exceed 2 EF points. Accordingly, when the axial shortening is included by using this formula, the variations in error due to K are virtually eliminated. Moreover, the spread of error values is markedly reduced over all EFs as evidenced by the virtually identical EF error curves that are close to the X-axis over the full range of EFs up to 65 – 70% for all values of K.

## DISCUSSION

This analysis demonstrates that the assumption of an ellipsoidal LV geometry at ED and ES leads to a simple structural formula for calculating the exact LVEF value that will be identical to the EF value by Simpson Rule planimetry, but requires only the radial and axial shortening fractions for input. No intermediate calculation of LV volume is necessary as in prior EF formulas such as Cube or Teicholz [6, 7]. These formulas rerquire converting the radial dimensions into the respective ED and ES volumes before calculating the EF.

Fractional shortening can be obtained from 2D or M-mode images. M-mode can be measured in parasternal and even in subcostal long and short axis views and often has superior temporal and spatial resolution than the simultaneous 2D image. It is not affected by frequently sub-optimal lateral and anterior wall endocardial resolution of apical views, and can be measured more precisely, and in multiple beats. Axial fractional shortening is measured in the apical view from the MV annular plane to the apical endocardium at ED and ES and also will not be affected by the variable resolution of the lateral and anterior walls in apical views that degrade planimetry.

Three linear approximations to the exact structural formula were investigated to further facilitate rapid and convenient clinical estimation of LVEF. Two of these formulas were single-factor, employing only the radial fractional shortening, while the third was a simple two-factor formula using both the radial and axial shortening to predict the exact EF. Optimal coefficient values for these formulas were obtained by comparing their EF results to the exact structural formula as the gold standard EF that is equivalent to planimetry in ventricles with ellipsoidal shape at ED and ES (appendix 1).

The simplicity of these formulas obviate computer assisted calculation and permit a “hand-held” quantitative measurement-based estimation of EF by the clinical reader. The analysis revealed that the errors in EF estimation will be less than **±** 8 to 10 EF points in almost all cases with any of the linear formulas and the average error would be smaller, depending on the distribution of K in the population. These error levels, if sustained in an empirical trial of these methods, compare favorably to that found by Pellikka et al [10], who reported a correlation of biplane echo EF with CMR EF of 0.66, echo EF results within 5% of CMR in only 43% of cases, and a mean absolute difference between biplane echo and CMR of 7.3% (N = 204). In Bland-Altman analysis of their data, 1.96 SDs in the difference between biplane echo EF and CMR were **−**15.2 to +20.1 EF percentage points. However, Pellikka included subjects with SWMA where errors may be higher than in uniformly contracting LVs and the time interval between the echo and CMR was not reported, both of which could have amplified their errors. In a meta-analysis of EF methods that included 174 studies (7047 subjects, Pickett et al [11] also reported a correlation of 0.66 between 2D echo and MRI but found narrower 1.96 SD limits than Pellikka, from **−**13.5 to +12.1 EF points in pooled Bland-Altman analysis.

The primary source of error in biplane echo EF is measurement error related to poor endocardial definition, particularly along the lateral and anterior walls in apical views where the echo beam is tangential to the surface. Fractional shortening methods are not as affected by this source of error so measurement error should be less significant. However, errors due to violation of the assumption of ellipsoidal shape at ED and ES may be significant and add to the error values shown in our purely geometric numerical simulation.

It is surprising and non-intuitive that simply doubling the radial fractional shortening could be used to estimate the LV EF. However, this analysis shows that the simplest formula EF ≈ 2F_D_, despite having the largest errors of the three linear approximations to exact LV EF, still appears to give reasonable estimates across a broad spectrum of clinically important EFs. Nonetheless there is a tendency for the one factor linear approximations to significantly overestimate EF at the highest EFs and for a more minimal overestimation at the very lowest EFs, the latter being clinically insignificant.

When the axial shortening was also included in the linear formula, a striking improvement in accuracy was evident (Figure 1, bottom panel). The numerical analysis also revealed that most of the remaining large errors consist of overestimation of EF at high values of radial fractional shortening (F_D_ > 35%) and associated high EFs above 65%. Awareness of this tendency to overestimation should help the clinical reader avoid systematic bias to overestimation in such cases. At lower EFs and radial fractional shortening values, which are of highest clinical importance, the EF errors are more symmetric and smaller in size.

Studies of global longitudinal strain (1) show that in LV dysfunction, reductions in longitudinal strain (F_L_) may be seen before EF changes and this is believed to be due to a compensatory increase in radial fractional shortening (F_D_) that preserves EF. Thus values of K (F_L_/ F_D_) will tend to be less than 40% in the majority of cases of LV dysfunction. These are represented by the upper curves in figure 1 (top and middle panels). A ratio above 100% represented by the lower curves would be seen in only a small minority of cases. Accordingly, curves for values of K of 80% or higher that result in a significant overestimation of EF by the linear approximations will be less likely to be seen in clinical cases than lower values.

The approximations developed here are not meant to replace the use of rigorous planimetry when it can be performed accurately, but to supplement it in cases in which it is technically difficult and there is concern as to its accuracy. This is not uncommon in non-contrast studies which are the routine method in outpatient and community hospital settings. It may also provide an objective adjunct to a visual assessment of LV function when planimetry is not possible at all. The planimetric EF should be within 8 - 10 points of the fractional shortening based one-factor linear approximation and within 5 - 6 points of the two-factor approximation. Other circumstances in which these linear estimations may be uniquely helpful are beat to beat variation in LV function such as in atrial fibrillation to obtain an average EF over several successive beats, and cases in which apical view end systolic foreshortening due to cardiac movement renders planimetry inaccurate, although model 3 would also not be accurate in the latter case. These simple linear formulas could be useful with the recent proliferation of POC echo performed with a variety of highly portable miniaturized echo devices in the ER or at the bedside by ER physicians, anesthesiologists, cardiologists and other clinicians. Finally, these linear models could be helpful for estimating EF in transesophageal echo (TEE) in the short axis sub-gastric view, either using the radial shortening alone or in conjunction with the deep gastric view.

Two other formulas have been proposed for non-planimetric calculation of EF from the radial LV dimension alone [6, 7]. The Cube formula uses a rigid assumption that the axial dimension is always twice the radial dimension. The Teicholz formula incorporates a correction factor based upon the radial dimension measurement that compensates for the average corresponding changes in the axial dimension that Teicholz et al measured on LV angiograms in patients undergoing cardiac catheterization. These formulas are more complex to calculate than either the linear or exact formula presented here and have not achieved wide use.

## LIMITATIONS

The formulas derived apply only to uniformly or nearly uniformly contracting ventricles, which comprise a substantial fraction of clinical cases, and should not be applied to cases with significant segmental wall motion abnormalities. Deviations from the exact EF that occur in the single factor linear models are due to variations in the axial to radial shortening ratio, K, that are not accounted for in models 1 and 2. Additional sources of error include deviation from ellipsoidal ED or ES shape despite apparent uniform contraction, and measurement errors of the radial and axial dimensions, but the latter source tends to be small.

We calculated and graphed the error as the difference of the approximate EF from the exact EF model in percentage points versus the exact EF. We did not use Bland Altman analysis [12], which would involve plotting the EF difference against the average of the two EFs because in our analysis the EF was not an empirical value subject to measurement error but rather an absolute and exactly correct value based on the assumed model.

Coefficients for Model 3 were derived by visual inspection, which introduced an element of subjectivity. However, this method allowed minimization of error in the clinically relevant range of EFs. Moreover, analytic methods cannot specify a unique set of coefficients when there are three unknown coefficients, as in the two-factor model, without applying constraints to the coefficient values, which would also introduce subjectivity.

Our study is a computational model-based simulation designed to explore the potential validity and performance of our novel approach to EF estimation. One advantage of using a simulation is that it is free from population sampling bias and the impact of technical measurement error and image quality. Simulation also allows potential pitfalls of the method to be identified. However, our findings, although promising, will require empirical data-based confirmation before clinical application.

## CONCLUSIONS

Geometric analysis shows that simple linear formulas based on LV fractional shortening have the potential to be used to estimate LV EF with reasonable accuracy in selected cases where planimetric assessment of LV EF is technically challenging or not feasible. The exact formula and the its two-factor linear approximation showed significantly smaller errors across the complete range of EFs and fractional shortening ratio variations, but were somewhat more difficult to apply than one factor formulas. Nonetheless, axial fractional shortening, though currently ignored, can be easily measured, and when combined with the radial fractional shortening in the formulas developed here, enables reliable assessment of LV EF that may be useful to clinical echocardiographers. Additional studies are needed to empirically validate these formulas.

## Data Availability

This study describes a computational (numerical) computer model of left ventricular contraction and does not contain any human subject or clinical data. All spreadsheets related to the model are available for review on request

## APPENDIX

## Appendix 1: Volume of An Arbitrarily Truncated Ellipsoid

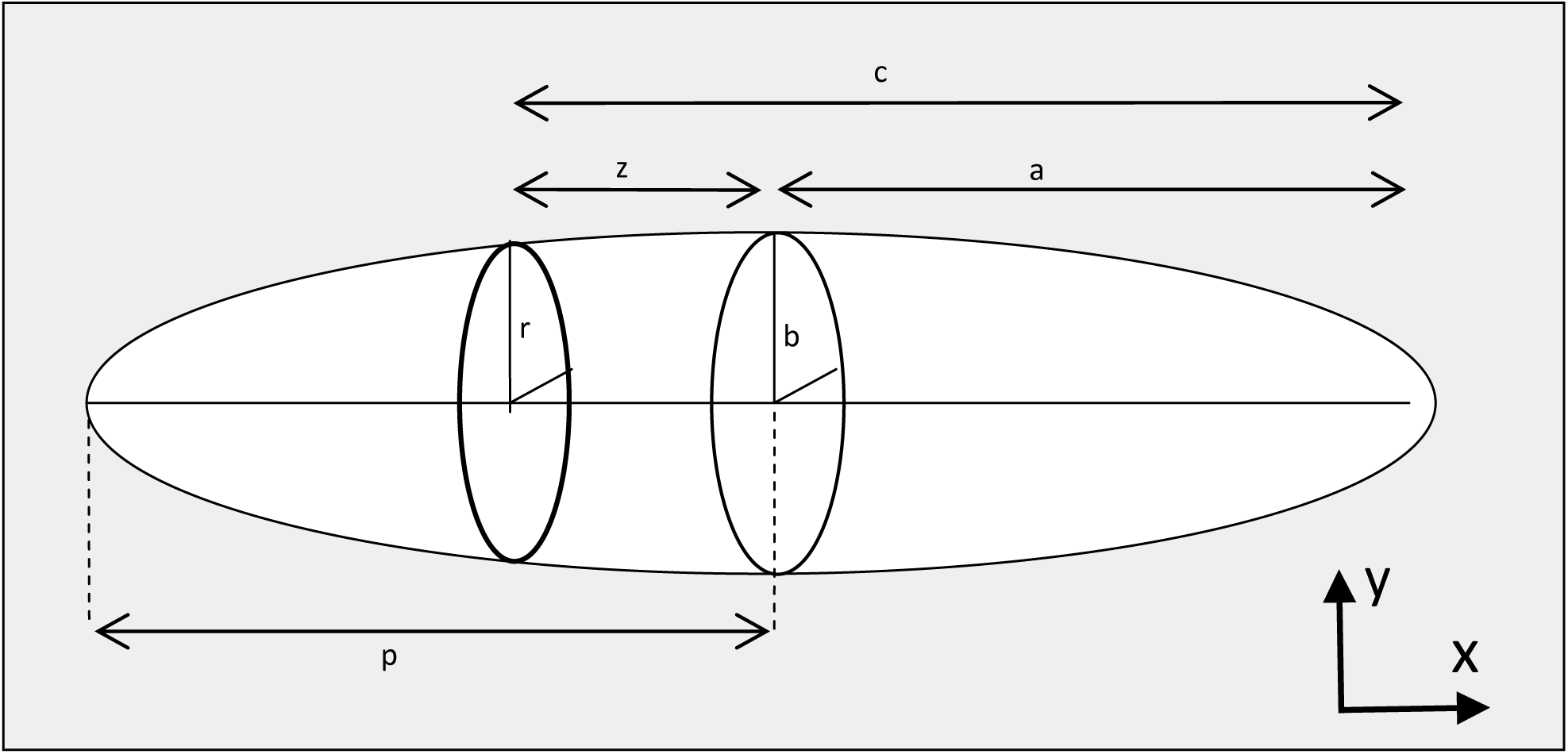

For a general ellipse in the x–y plane 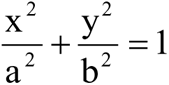, and 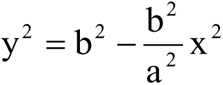

For a generalized prolate ellipsoid obtained by rotation of the ellipse around the x axis:

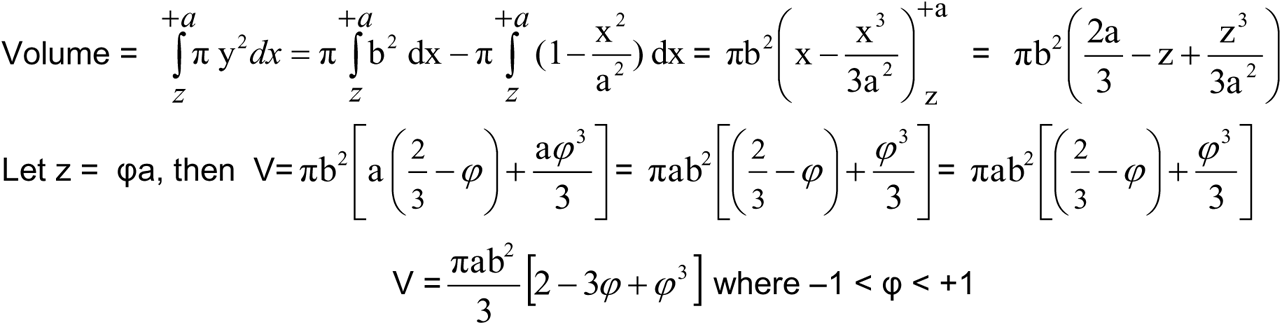

Thus, for any arbitrarily truncated ellipse, the volume remains a constant term multiplied by πab^2^, and changes in radius and length alone do not alter the constant factor.

The following are familiar special cases of this result.

Case 1: For or *φ* =0, V = (2/3)πab^2^, the familiar formula for a hemiellipse.

Case 2: For *φ* = -1, V = (4/3)πab^2^, the familiar formula for a complete ellipse.

Note that for *φ* = **+**1, V = 0 as expected.

## Appendix 2: Exact Formula for EF

Using case #1 in appendix 1 above, the volume of a hemiellipse = (4/3)π(D/2)^2^ L where D = ellipse diameter or LV diameter at the base and L = axial length. The ejection fraction is then given by:

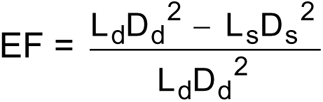

where d and s denote diastole and systole and the constants have cancelled out. Dividing numerator and denominator by 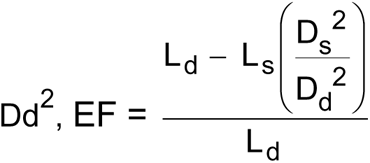, and then dividing by Ld,

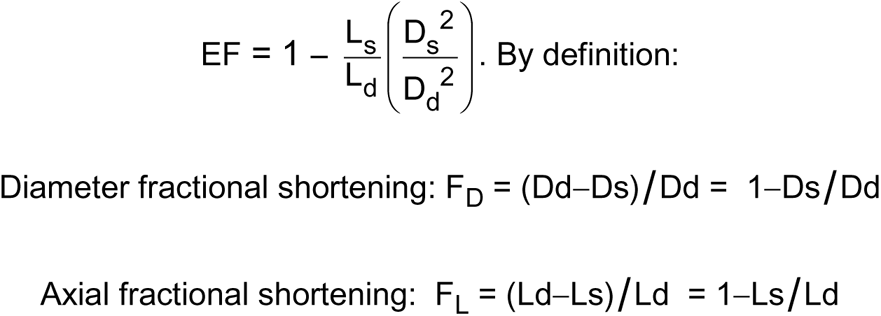

Substituting (1−F_D_)^2^ = (Ds/Dd)^2^ and (1−F_L_) = Ls/Ld in the preceding expression for EF yields:

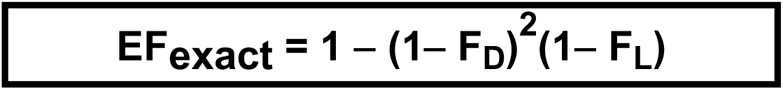

## Appendix 3

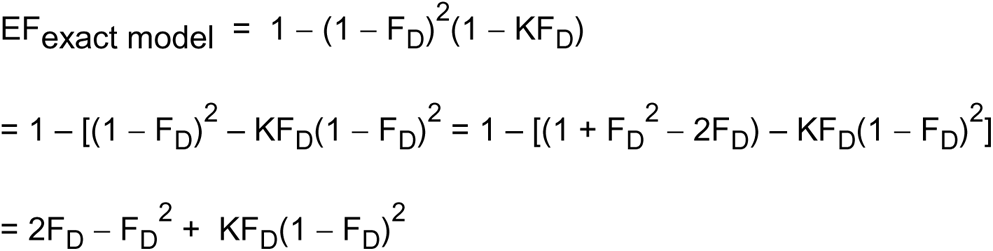

